# Health and economic outcomes of newborn screening for infantile-onset Pompe disease

**DOI:** 10.1101/2020.04.28.20080606

**Authors:** John S. Richardson, Alex R. Kemper, Scott D. Grosse, Wendy K.K. Lam, Angela M. Rose, Ayesha Ahmad, Achamyeleh Gebremariam, Lisa A. Prosser

**Author notes:** Corresponding Author: Lisa A. Prosser, 300 North Ingalls St. 6D04, Ann Arbor, MI 48109.

## Abstract

**Purpose:** To estimate health and economic outcomes associated with NBS for infantile-onset Pompe disease in the United States.

**Methods:** A decision analytic microsimulation model simulated health and economic outcomes of a birth cohort of 4 million children in the United States. Universal NBS and treatment was compared to clinical identification and treatment of infantile-onset Pompe disease. Main outcomes were projected cases identified, costs, quality adjusted life-years (QALYs), and incremental cost-effectiveness ratios (ICERs) over the life course.

**Results:** Universal NBS for Pompe disease and confirmatory testing was estimated to cost an additional $26 million annually. Additional medication costs associated with earlier treatment initiation were $181 million; however, $8 million in medical care costs for other services were averted due to delayed disease progression. Infants with screened and treated infantile-onset Pompe disease experienced an average lifetime increase of 11.66 QALYs compared to clinical detection. The ICER was $408,000/QALY from the health care perspective and $379,000/QALY from a societal perspective. Results were sensitive to the cost of enzyme replacement therapy.

**Conclusions:** Newborn screening for Pompe disease results in substantial health gains for individuals with infantile-onset Pompe disease, but with additional costs.

## Introduction

State-based newborn screening (NBS) programs have expanded over the past 50 years from screening for phenylketonuria to screening for more than 30 conditions.^1,2^ The Advisory Committee on Heritable Disorders in Newborns and Children (ACHDNC) was developed to support state NBS decisions by reviewing proposed conditions and synthesizing evidence regarding the feasibility and benefits of screening.^3^ The ACHDNC makes recommendations to the Secretary of Health and Human Services, who in turn determines a Recommended Uniform Screening Panel (RUSP); cost-effectiveness is not a criterion in the recommendation process.^4^

Pompe disease was recommended by the ACHDNC in June 2013 and was added to the RUSP in March 2015.^5^ It is a rare condition that is identified in about 1 in 40,000 births.^6^ Pompe disease occurs from a defect in the *GAA* gene leading to the accumulation of lysosomal glycogen and, depending on the form and severity, can result in cardiomyopathy, progressive muscle weakness, respiratory failure, and heart failure.^7^ Pompe disease can be treated with alglucosidase alfa, an enzyme replacement therapy (ERT) that costs between $250,000 and $500,000 per year for biweekly infusions.^8^

The purpose of this study is to evaluate the cost-effectiveness of universal NBS for infantile-onset Pompe disease compared to usual clinical identification.

## Materials and Methods

### Study Design and Scope

We created a decision analytic microsimulation model for a US cohort of 4 million newborns.^9^ Microsimulations are mathematical models that track the unique health trajectory and health outcomes of each simulated person over time. The model compared outcomes for two strategies: (1) clinical identification of infantile-onset Pompe disease and treatment with ERT; and (2) NBS for Pompe disease and treatment of infantile-onset disease with ERT. The analytic time horizon included the life-course of the birth cohort. We used one-year time increments to capture changes in health status over each individual’s life. We incorporated costs and outcomes from the healthcare sector and societal perspectives.^10^ Insufficient data are available to model the long-term costs and outcomes for individuals identified through NBS with late-onset Pompe disease.

### Model Structure and Assumptions

In the clinical identification scenario, individuals were diagnosed with Pompe disease at varying ages based on testing prompted by the recognition of symptoms. This analysis used a classification of Pompe disease developed for the ACHDNC evidence review.^4,11^ The cases were classified as infantile-onset (those with onset before age 12 months) or late-onset. Infantile-onset cases were further classified as either with or without cardiomyopathy (Figure 1). A more recent set of guidelines defines infantile-onset as only those with cardiomyopathy and onset before 12 months; however, these were published after our analytic framework had been defined.^12^ Once identified as having Pompe disease, an individual in our analysis can remain at the current stage of disease severity, progress to later stages of symptoms or die.

**Figure 1.**
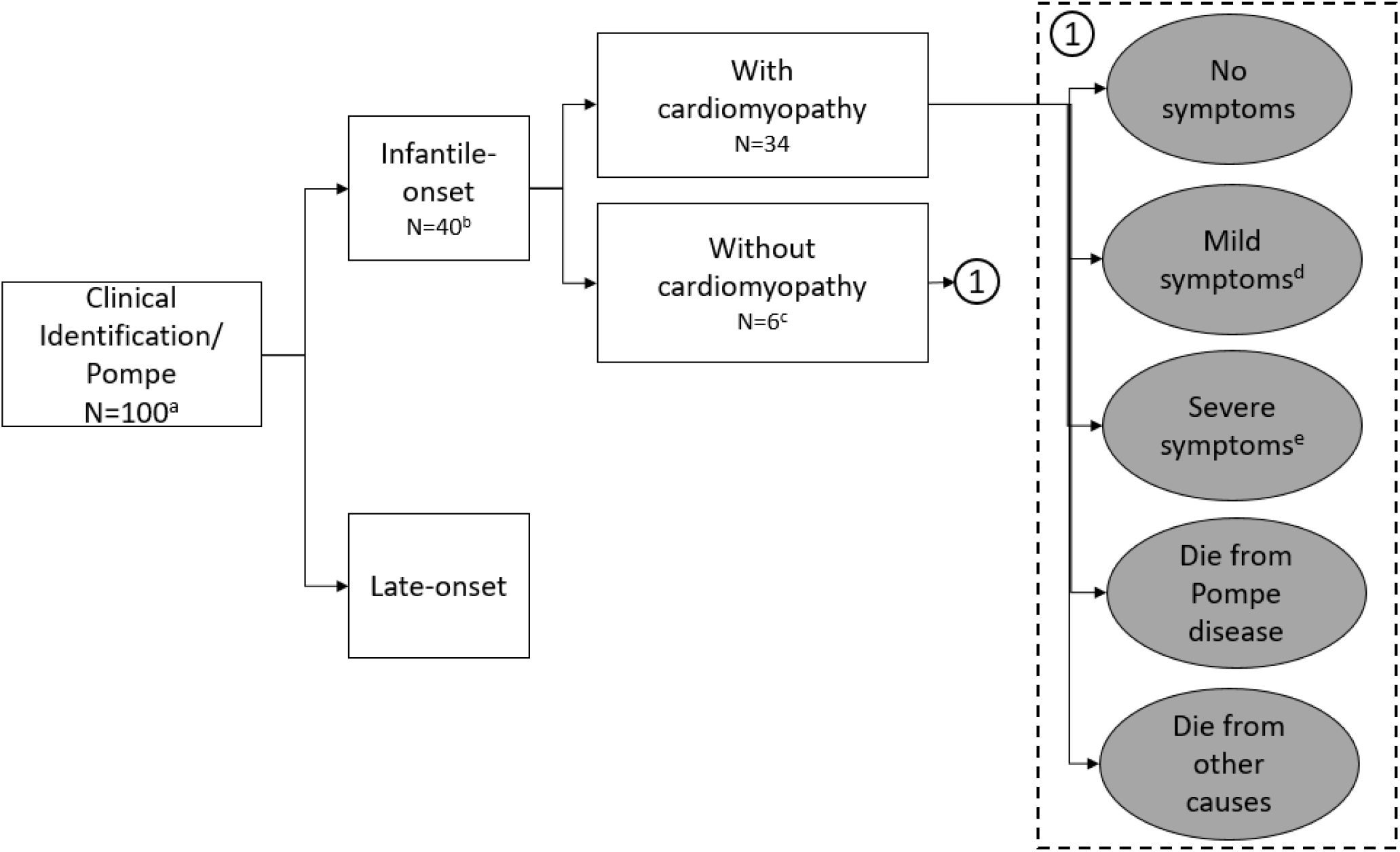
Model framework. **a**. **Clinical identification** ^a^ Cohort of 4,000,000 ^b^ Infantile-onset Pompe disease was defined as symptom onset prior to 12 months. ^c^ Two-thirds of patients identified with infantile-onset Pompe disease without cardiomyopathy experience a delay in treatment and do not start treatment until age 3 ^d^ Muscle weakness and declining muscle tone, but maintain the ability to walk and perform most activities independently ^e^ Increasing muscle weakness and breathing problems necessitating wheelchair use and ventilator dependence **b**. **Newborn screening** ^a^ Infantile-onset Pompe disease was defined as symptom onset prior to 12 months. ^b^ Muscle weakness and declining muscle tone, but maintain the ability to walk and perform most activities independently ^c^ Increasing muscle weakness and breathing problems necessitating wheelchair use and ventilator dependence ^d^ Since the focus of this study is on screening for and the identification of infantile-onset Pompe, we have not distinguished the potential reasons for false positives such as being a carrier or other technical reasons ^e^6 of the 10 false negatives are expected to have a pseudodeficiency and never develop any symptoms

In the NBS scenario, we assume that newborns are screened within 2 days of birth. If a newborn has a positive screen, a repeat test is conducted, followed by, if indicated, confirmatory testing of enzyme activity and molecular testing and diagnosis by 2-3 weeks of age. Infants with low enzyme activity and *GAA* variants consistent with pseudodeficiency alleles are not evaluated further. Individuals diagnosed with Pompe disease are classified as infantile-onset (with or without cardiomyopathy) or probable late-onset depending on whether signs or symptoms are present during confirmatory testing. Those in the NBS scenario with infantile-onset Pompe progress through the same set of long-term health states as those in the clinical identification scenario. Probability of a false negative result is also included in the NBS scenario. It is assumed that all such cases would be late-onset.

The focus of our analysis is on infantile-onset cases and we did not evaluate changes in health outcomes or costs of treatment for late-onset Pompe disease. However, we did include an extra annual physician visit for asymptomatic patients screened and diagnosed with probable Pompe disease who had not yet exhibited symptoms as standard of care.

In the clinical identification scenario we assumed that identification of infantile-onset Pompe disease among those with cardiomyopathy occurred at 4.5 months of life.^6^ In the absence of screening, we assumed that one-third of those with infantile-onset Pompe disease but no cardiomyopathy were identified and initiated treatment earlier than one year, while two-thirds were identified after the first year of life with an average age of diagnosis at three years before being identified and starting treatment.

The life-course trajectory of infantile-onset Pompe disease was captured using a state-transition simulation model with the following states: no symptoms, mild symptoms, severe symptoms, died from Pompe disease, and died from other causes (Figure 1). This state-transition model was a part of the microsimulation model and tracks individuals as they cycle through the different health states over consecutive 1-year time periods. Mild symptoms included some signs and symptoms. Ventilator dependence is required for the classification of severe disease.

Model inputs were derived using published reports, unpublished data, and judgements from an expert panel (Table 1). The expert panel consisted of six individuals (see the Acknowledgements section) with extensive experience in newborn screening, studying rare genetic conditions, and treating individuals with Pompe disease. The parameters determined from the expert panel were developed using a modified Delphi or “Decision Delphi” approach.^13^

**Table 1.**
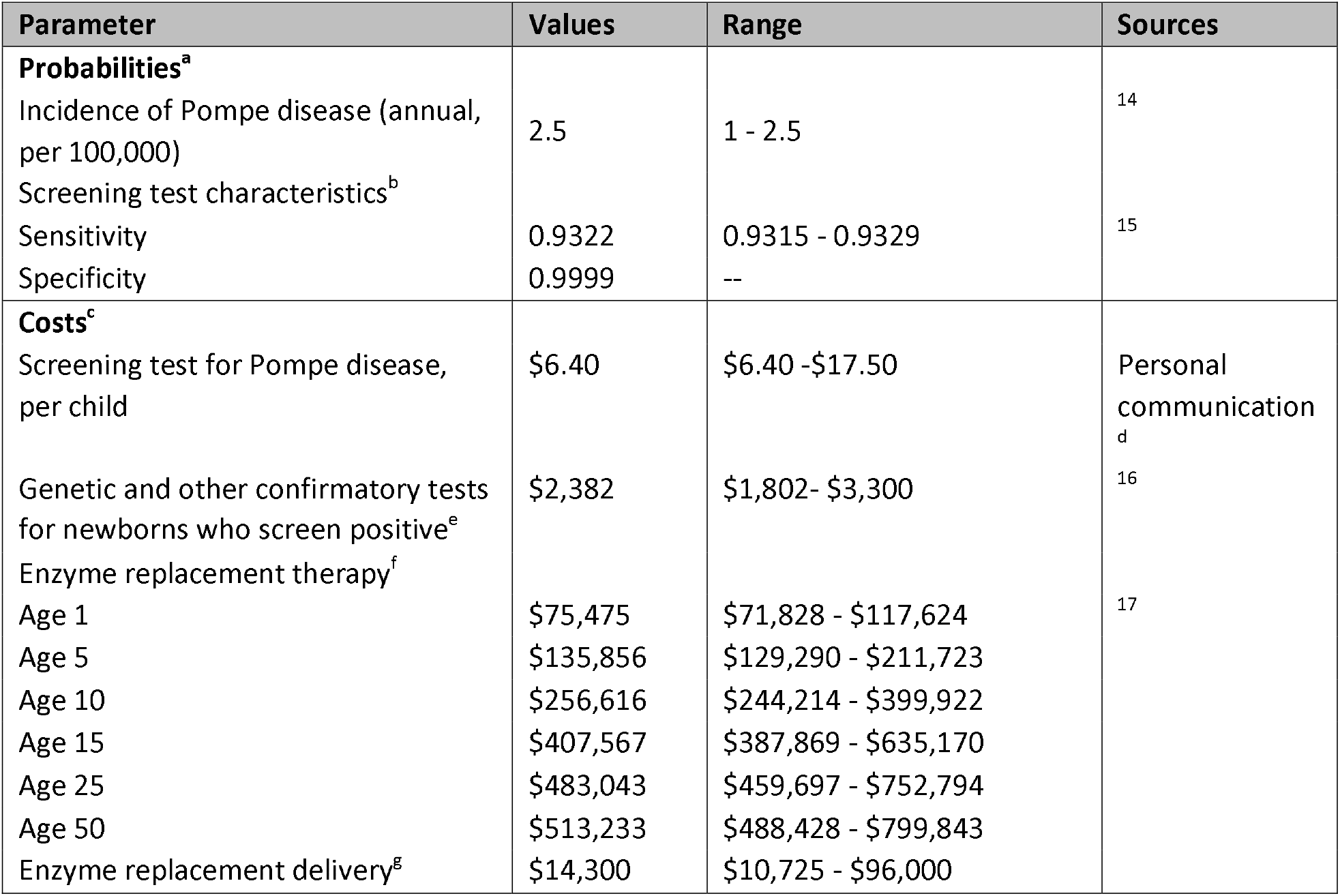
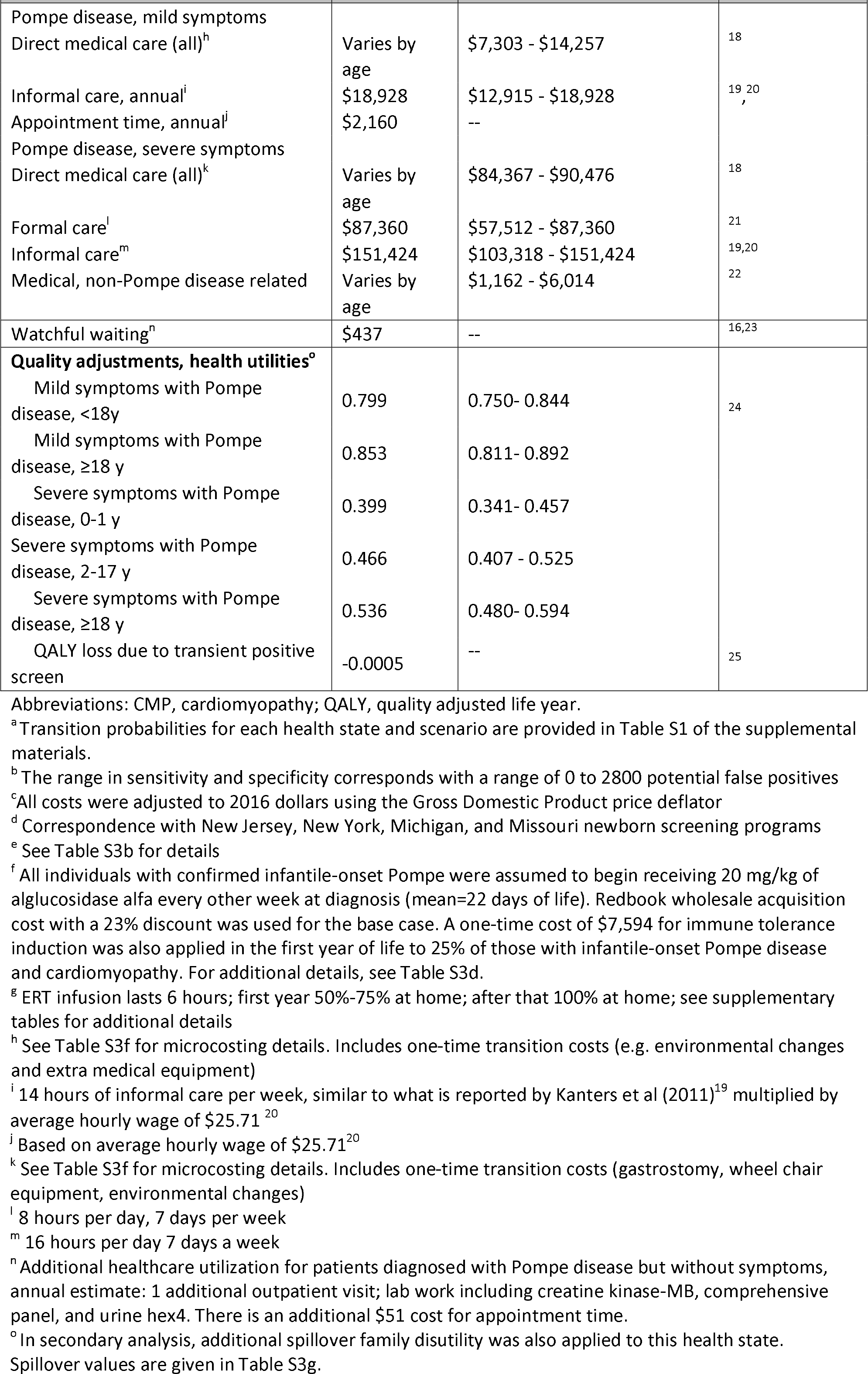
Model inputs.

| Parameter | Values | Range | Sources |
| --- | --- | --- | --- |
| **Probabilities<sup>a</sup>** |  |  |  |
| Incidence of Pompe disease (annual, per 100,000) | 2.5 | 1 - 2.5 | 14 |
| Screening test characteristics<sup>b</sup> |  |  |  |
| Sensitivity | 0.9322 | 0.9315 - 0.9329 | 15 |
| Specificity | 0.9999 | -- |  |
| **Costs<sup>c</sup>** |  |  |  |
| Screening test for Pompe disease, per child | \$6.40 | \$6.40 - \$17.50 | Personal communication<sup>d</sup> |
| Genetic and other confirmatory tests for newborns who screen positive<sup>e</sup> | \$2,382 | \$1,802- \$3,300 | 16 |
| Enzyme replacement therapy<sup>f</sup> |  |  |  |
| Age 1 | \$75,475 | \$71,828 - \$117,624 | 17 |
| Age 5 | \$135,856 | \$129,290 - \$211,723 | |
| Age 10 | \$256,616 | \$244,214 - \$399,922 | |
| Age 15 | \$407,567 | \$387,869 - \$635,170 | |
| Age 25 | \$483,043 | \$459,697 - \$752,794 | |
| Age 50 | \$513,233 | \$488,428 - \$799,843 | |
| Enzyme replacement delivery<sup>g</sup> | \$14,300 | \$10,725 - \$96,000 | |
| Pompe disease, mild symptoms<br>Direct medical care (all)<sup>h</sup> | Varies by age | \$7,303 - \$14,257 | 18 |
| Informal care, annual<sup>i</sup> | \$18,928 | \$12,915 - \$18,928 | 19, 20 |
| Appointment time, annual<sup>j</sup> | \$2,160 | -- | |
| Pompe disease, severe symptoms<br>Direct medical care (all)<sup>k</sup> | Varies by age | \$84,367 - \$90,476 | 18 |
| Formal care<sup>l</sup> | \$87,360 | \$57,512 - \$87,360 | 21 |
| Informal care<sup>m</sup> | \$151,424 | \$103,318 - \$151,424 | 19,20 |
| Medical, non-Pompe disease related | Varies by age | \$1,162 - \$6,014 | 22 |
| Watchful waiting<sup>n</sup> | \$437 | -- | 16,23 |
| **Quality adjustments, health utilities<sup>o</sup>** |  |  |  |
| Mild symptoms with Pompe disease, <18y | 0.799 | 0.750- 0.844 | 24 |
| Mild symptoms with Pompe disease, ≥18 y | 0.853 | 0.811- 0.892 |  |
| Severe symptoms with Pompe disease, 0-1 y | 0.399 | 0.341- 0.457 |  |
| Severe symptoms with Pompe disease, 2-17 y | 0.466 | 0.407 - 0.525 |  |
| Severe symptoms with Pompe disease, ≥18 y | 0.536 | 0.480- 0.594 |  |
| QALY loss due to transient positive screen | -0.0005 | -- | 25 |

Abbreviations: CMP, cardiomyopathy; QALY, quality adjusted life year.
<sup>a</sup> Transition probabilities for each health state and scenario are provided in Table S1 of the supplemental materials.
<sup>b</sup> The range in sensitivity and specificity corresponds with a range of 0 to 2800 potential false positives
<sup>c</sup> All costs were adjusted to 2016 dollars using the Gross Domestic Product price deflator
<sup>d</sup> Correspondence with New Jersey, New York, Michigan, and Missouri newborn screening programs
<sup>e</sup> See Table S3b for details
<sup>f</sup> All individuals with confirmed infantile-onset Pompe were assumed to begin receiving 20 mg/kg of alglucosidase alfa every other week at diagnosis (mean=22 days of life). Redbook wholesale acquisition cost with a 23% discount was used for the base case. A one-time cost of \$7,594 for immune tolerance induction was also applied in the first year of life to 25% of those with infantile-onset Pompe disease and cardiomyopathy. For additional details, see Table S3d.
<sup>g</sup> ERT infusion lasts 6 hours; first year 50%-75% at home; after that 100% at home; see supplementary tables for additional details
<sup>h</sup> See Table S3f for microcosting details. Includes one-time transition costs (e.g. environmental changes and extra medical equipment)
<sup>i</sup> 14 hours of informal care per week, similar to what is reported by Kanters et al (2011)<sup>19</sup> multiplied by
average hourly wage of \$25.71<sup>20</sup>
<sup>j</sup> Based on average hourly wage of \$25.71<sup>20</sup>
<sup>k</sup> See Table S3f for microcosting details. Includes one-time transition costs (gastrostomy, wheel chair equipment, environmental changes)
<sup>l</sup> 8 hours per day, 7 days per week

**Table 2.** Projected health and economic outcomes.

| Outcomes | Clinically Diagnosed & Treated | Newborn Screening & Treated |
| --- | --- | --- |
| **Health outcomes, infantile-onset** |  |  |
| Screening outcomes (or within <u>first year</u> of life for clinically diagnosed) |  |  |
| Cases, infantile-onset with cardiomyopathy | 34 | 34 |
| Cases, infantile-onset without cardiomyopathy<sup>a</sup> | 2 | 6 |
| Proportions with cardiomyopathy, by health state, at age 10 |  |  |
| No symptoms | 0.00 | 0.30 |
| Mild symptoms | 0.19 | 0.67 |
| Severe symptoms | 0.10 | 0.00 |
| Dead from Pompe disease | 0.70 | 0.02 |
| Dead from other causes | 0.01 | 0.01 |
| Health outcomes |  |  |
| By age 3 |  |  |
| Ventilator dependent, n | 7.11 | 0.53 |
| Deaths, n | 6.93 | 0.00 |
| By age 10 |  |  |
| Ventilator dependent, n | 4.87 | 0.17 |
| Deaths, n | 11.58 | 0.00 |
| By age 20 |  |  |
| Ventilator dependent, n | 4.06 | 0.32 |
| Deaths, n | 14.40 | 0.02 |
| **Disaggregated results among individuals diagnosed with infantile-onset Pompe disease (per person)<sup>b</sup>** |  |  |
| Screening and confirmation costs | NA | \$2,388 |
| Enzyme replacement therapy (ERT) | \$6,462,230 | \$10,985,209 |
| Medical, Pompe disease other than ERT | \$385,039 | \$178,834 |
| Medical, non-Pompe disease | \$40,731 | \$72,952 |
| Formal caregiving | \$283,564 | \$26,534 |
| Informal caregiving | \$774,734 | \$428,162 |
| Non-medical (e.g. appointment time costs and environmental modifications) | \$33,318 | \$44,607 |
| Total costs, healthcare sector<sup>c</sup> | \$7,171,563 | \$11,265,916 |
| Total costs, societal | \$7,979,615 | \$11,738,685 |
| Mean QALYs | 15.01 | 26.67 |
| Infantile-onset with cardiomyopathy | 13.76 | 26.65 |
| Infantile-onset without cardiomyopathy | 22.11 | 26.79 |

| **Outcomes** | **Clinically Diagnosed & Treated** | **Newborn Screening & Treated** |
| --- | --- | --- |
| **Summary and disaggregated results for annual US newborn cohort (n=4,000,000)<sup>e</sup>** |  |  |
| Costs |  |  |
| Screening and confirmation tests | \$0 | \$26,224,278 |
| Enzyme replacement therapy | \$258,489,185 | \$439,408,352 |
| Pompe disease-related medical care | \$15,747,528 | \$7,639,698 |
| Non Pompe disease-related medical care | \$291,659,614,973 | \$291,660,903,800 |
| Formal caregiving costs | \$11,342,569 | \$1,061,343 |
| Informal caregiving | \$30,989,365 | \$17,126,462 |
| Non-medical (e.g., appointment time costs and environmental modifications) | \$1,875,353 | \$2,355,840 |
| Total costs (healthcare sector<sup>f</sup>) | \$291,945,194,255 | \$292,135,237,471 |
| Total costs (societal) | \$291,978,058,973 | \$292,154,719,773 |
| Incremental costs (healthcare sector) | -- | \$190,043,216 |
| Incremental costs (societal) | -- | \$176,660,800 |
| Total QALYs | 121,806,506 | 121,806,973 |
| Incremental QALYs | -- | 466 |
| ICER<sup>g</sup> (healthcare sector) | -- | \$408,000/QALY |
| ICER<sup>g</sup> (societal) | -- | \$379,000/QALY |

Abbreviation: ICER=incremental cost-effectiveness ratio.
<sup>a</sup> Under clinical identification, 4 of the 6 cases are anticipated to remain undiagnosed until age 3.
<sup>b</sup> Average per person costs and QALYs. Costs and QALYs discounted at 3%.
<sup>c</sup> Excludes informal caregiving and non-medical costs
<sup>d</sup> Weighted average of two cases treated near birth and four cases treated at age three
<sup>e</sup> Costs and QALYs are discounted at 3% to 2016 USD.
<sup>f</sup> The healthcare sector perspective includes all but non-medical and informal caregiving costs.
<sup>g</sup> ICERs were calculated using additional decimal places for incremental QALYs – see supplementary tables for detailed results.

### Epidemiology Inputs

The sensitivity and specificity of NBS as well as other screening probabilities were derived from a pilot screening program in Taiwan and confirmed by an expert panel (Table S2).^15^ Transition probabilities were calibrated to short-term health outcomes (to age 3) using published and unpublished data and longer-term outcome trajectories determined by an expert panel.^26,27^ (Figure S2, Table S1) Based on input from the expert panel, we also assumed that early treated individuals with infantile-onset Pompe disease have the same underlying risk of neurological or other conditions as the general pediatric population.

### Costs

Costs included the costs of screening all newborns, doing confirmatory testing among positive screens, and the costs associated with treatment for patients diagnosed with infantile-onset Pompe disease. Both medical costs and time costs for informal caregivers (for the societal perspective analysis) were included (Table S3). All costs were adjusted to 2016 dollars (Table S3). The cost of screening was determined through key informant interviews with U.S. states that were either in the process of starting to implement or had established screening for Pompe disease. A base case estimate of $6.40 per newborn was used for the cost of the initial screen (Table 1). Our cost estimates for false positive screens presume that programs use an assay with specificity comparable to that reported by the New York screening program rather than one of the less efficient assays.^28^ In the base case, we assumed full adherence to treatment recommendations and that no family would deny treatment for their child. Patients with confirmed infantile-onset Pompe disease were assumed to receive an infusion of 20 mg/kg every other week, resulting in medication costs that ranged from an estimated $75,475 to $513,233 per year depending on age and on the assumed cost of the drug in addition to administration costs of $14,300 per year.

We used a micro-costing approach to estimate the total costs of care associated with each health state (Table S3). A micro-costing approach establishes estimates for each of the components of resource units and then aggregates them to get the overall costs. In addition to the costs of ERT, ongoing costs of care included costs of additional required medications, specialist visits, emergency visits, hospitalizations, laboratory tests, and medical equipment, as well as formal and informal caregiving (Table S3). The costs of these medical services were based on the Centers for Medicare and Medicaid Services Physician Fee Schedule, and the number of services were based on estimates from clinical experts (Table S3). Informal caregiving is care provided by a family member or other individual who is not paid for his or her time. Informal caregiving hours were assumed to range from 14 hours per week for patients with few symptoms to 16 hours per day for severe forms.^19^ Formal caregiving hours were assumed to be 8 hours per day for those with severe symptoms. Medical expenditures unrelated to Pompe disease were also included.^29^

### Quality of Life

Health utility weights for each health state were determined from a separate study using time trade-off questions fielded to a national community-based sample.^30^ The values for those with mild symptoms ranged from 0.799 to 0.853, and for those with severe symptoms ranged from 0.399 to 0.536, depending on age. In secondary analyses, we also included reductions in health utility for family members attributable to the patient’s illness (family spillover utility) as well as a small and transient reduction in health utility associated with the experience of a false positive screen.

## Analysis Plan

The primary outcome is the incremental cost-effectiveness ratio (ICER) in dollars per quality-adjusted life year gained for the NBS scenario relative to the clinical identification scenario. Incremental costs include the costs of screening, short-term follow-up, and diagnostic testing for all infants who screen positive. Secondary outcomes included numbers of cases identified with and without screening, disaggregated outcomes (i.e., the major components contributing to the numerator and denominator of the ICER), and undiscounted outcomes (i.e., the outcomes in 30 years are assumed to have the same value as outcomes over the next year). The base case analysis used most likely point estimates for parameters, and included test characteristics and screening costs derived from the New York screening program. We conducted one-way sensitivity analyses of screening parameters, transition probabilities, and cost parameters to determine the effect of varying individual parameters within a plausible range on the incremental cost-effectiveness ratio. Given that clinicians sometimes increase the dosage or frequency of ERT, we examined secondary scenarios of 40 mg per kg every other week and every week.^31^ Scenario analyses were also conducted for a broader range of hypothetical reduction in the cost of ERT, the inclusion of family spillover effects on quality of life, and a comparison between clinical identification with and without ERT. Costs and quality-adjusted life years (QALYs) were discounted using a 3% annual discount rate. Simulation software was TreeAge Pro 2017 version R2.1 (TreeAge Software, Inc., Williamstown, MA).

**Code Availability**The model is available upon request by contacting the corresponding author.

## Results

### Screening Test Outcomes and Diagnoses

Using base case parameter input values, 100 individuals in the simulated birth cohort would be identified with Pompe disease in both scenarios. Among these individuals, 34 have infantile-onset Pompe disease with cardiomyopathy, 6 infantile-onset (atypical) Pompe disease without cardiomyopathy, and 60 have late-onset Pompe disease. Newborn screening is also assumed to lead to the identification of 38 individuals who have low alpha-glucosidase enzyme activity due to carrier status or a pseudodeficiency in a *GAA* allele that does not cause clinical symptoms.

### Health Outcomes

Newborn screening resulted in substantial health gains as measured by the proportion of patients who are ventilator-dependent, Pompe-disease related deaths, and QALYs for those with infantile-onset Pompe disease (Table 2). The QALYs gained from NBS compared to clinical identification was 13 QALYs per person with infantile-onset Pompe disease with cardiomyopathy. For infantile-onset Pompe disease without cardiomyopathy (atypical presentation), the gain was 4.7 QALYs per person. For a cohort of 4 million children, NBS results in a cumulative lifetime gain of 466 QALYs compared to clinical identification.

### Costs

The total discounted lifetime costs of care for infantile-onset Pompe disease were projected at $7.98 million for each person who is clinically identified (Table 2). For infantile-onset Pompe disease identified through screening, average lifetime costs were estimated to be $11.7 million. Increases in costs of care associated with the screened group were predominantly from additional time receiving ERT due to earlier initiation of treatment combined with longer overall period of treatment due to averted deaths. Increased treatment costs were only slightly offset by improved health, resulting in lower infantile-onset Pompe disease-related costs other than ERT (-$0.2 million), formal caregiving costs (-$0.26 million) and informal caregiving costs (-$0.35 million).

At the population level, NBS and confirmatory tests for 4 million births were estimated to cost $26 million (Table 2). Total incremental costs associated with NBS strategy for infantile-onset Pompe disease were $177 million from the societal perspective and $190 million from the health sector perspective. ERT costs were estimated to increase by $181 million under NBS. The category of cost with the largest decrease attributable to NBS was informal caregiving costs ($14 million).

### Cost-effectiveness

Using base case assumptions, the ICER for NBS compared with clinical identification of infantile-onset Pompe disease was $379,000/QALY from a societal perspective and $408,000/QALY from a health sector perspective (Table 2). If the cost of screening were $17.50 per test rather than $6.40 per test, NBS would cost society 25% more or $474,000/QALY. Lower estimates for the cost of ERT resulted in a cost-effectiveness ratio of $303,000/QALY and higher estimates resulted in $402,000/QALY (Figure 2, Table S4). Varying treatment effectiveness across the plausible range yields cost-effectiveness ratios from $311,000/QALY to $420,000.

**Figure 2.**
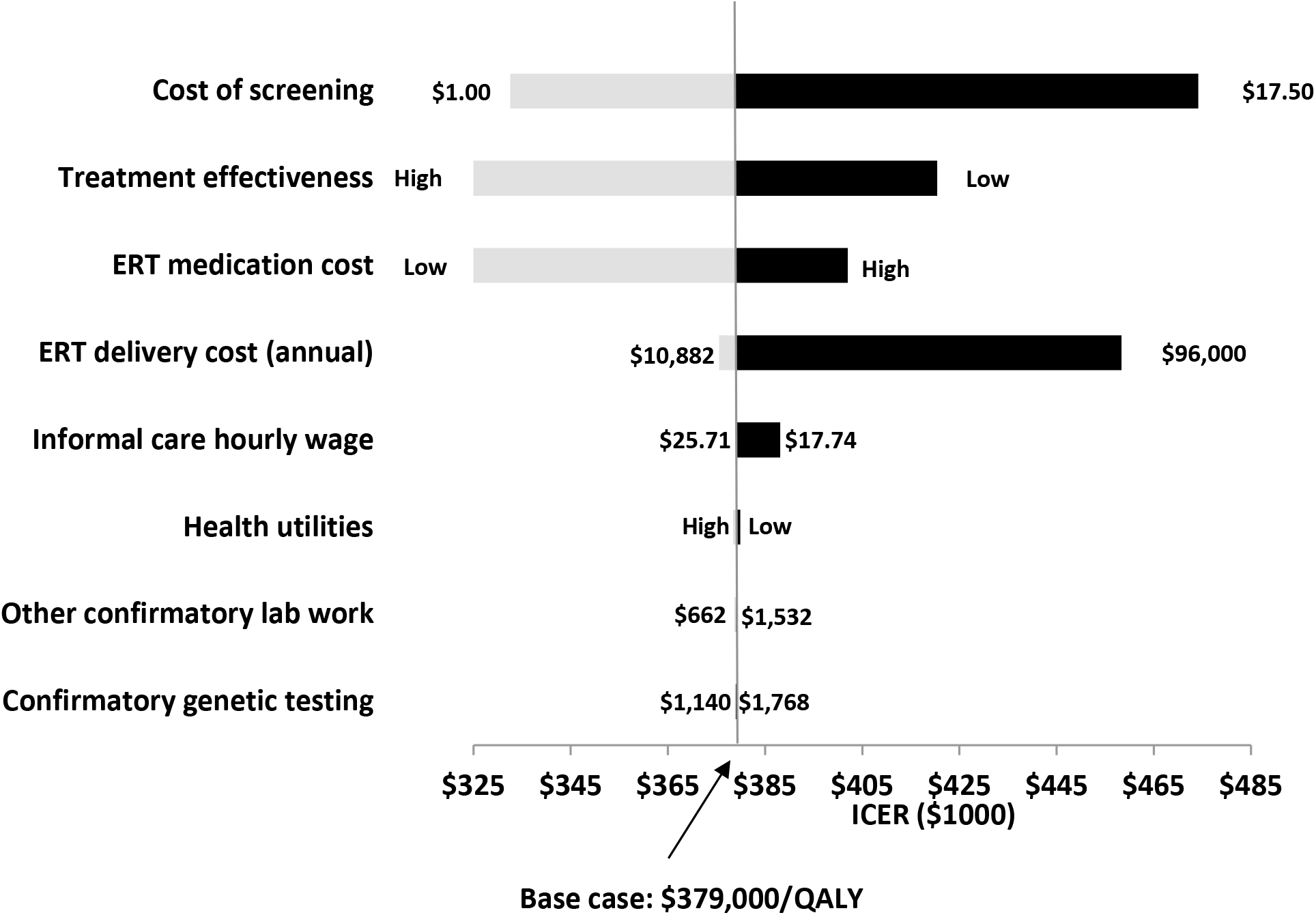
One-way sensitivity analysis.

Figure S1 and Table S5 shows a scenario analysis of reductions in the cost per dose of alglucosidase alfa. In scenario analysis, if the cost of the drug were reduced by 80% (discount to average wholesale price), the cost-effectiveness ratio would be approximately $102,000 /QALY (Figure S1 and Table S5). Results were robust to changes in other parameters. Including a family spillover utility for one parent yielded slightly more favorable cost-effectiveness results of $372,000/QALY gained (Table S6). Increasing the dosage of ERT to 40 mg per kg every other week or every week substantially increases the ICER to $750,000/QALY to $1,500,000/QALY (Table S7).

In a secondary analysis, we evaluated the cost-effectiveness of ERT by comparing outcomes of clinical identification with ERT to clinical identification without ERT (in the absence of newborn screening). This analysis yields cost-effectiveness ratios greater than $500,000/QALY for clinical identification and treatment of infantile-onset Pompe disease (Table S8).

### Discussion

We found that NBS for Pompe disease in the United States would provide substantial gains in health and quality of life among individuals with infantile-onset Pompe disease. This comes, however, with increased costs due to earlier initiation of treatment and extended lifespan for individuals with infantile-onset Pompe disease. We measured the value gained from these increased costs through cost-effectiveness analysis.^32,33^ In situations where resources are limited, unofficial thresholds for ICERs are sometimes used in the United States to encourage investment in high value or low cost per QALY services. An ICER of $379,000/QALY is far beyond the upper bound of the unofficial range of commonly used cost-effectiveness benchmarks ($50,000$150,000/QALY) for the United States.^34^ Some, however, argue that different criteria should be applied for the cost-effectiveness of treatments for rare conditions. The UK National Institute for Health and Clinical Excellence (NICE) for ‘highly specialized technologies’ for conditions with a prevalence of 2 per 100,000 or less sets a cost-effectiveness threshold that is roughly 3 times higher than the general threshold.^35,36^ If a similar approach were applied in the United States to interventions for very rare disorders (i.e., a threshold range of $150,000 to $450,000/QALY), screening for infantile-onset Pompe disease would fall at the very upper end of what could be considered cost-effective using these alternate criteria. Other countries also cite additional considerations for treatments for rare diseases but have not put forth an adjusted threshold.^35^ If higher dosages and frequency of ERT are delivered than the base case scenario, then newborn screening for infantile-onset Pompe disease would not meet thresholds for cost-effectiveness even using the most liberal criteria for assessing the cost-effectiveness of rare conditions.

This analysis extends previous work conducted on behalf of the Condition Review Workgroup for the Advisory Committee on Heritable Disorders in Newborns and Children.^4,6^ It evaluates the cost-effectiveness of NBS for infantile-onset Pompe disease, including costs and outcomes throughout the lifespan. In a prior analysis, we estimated that NBS in the United States would prevent 13 deaths and 26 people from needing mechanical ventilation by age 3 years. The current analysis extends these results into a lifetime simulation model to answer key questions on lifetime health and quality-of-life benefits while also incorporating screening and disease-related costs.

This analysis uses the strategy of clinical identification followed by treatment with alglucosidase alfa as the primary comparator strategy, recognizing that individuals identified clinically are being offered and typically will initiate treatment with alglucosidase alfa. In a secondary analysis comparing clinical identification with ERT to clinical identification without ERT, we estimated a much higher cost-effectiveness ratio. Even if higher thresholds are considered, this leaves us in a challenging position in which NBS for a condition might potentially be considered cost-effective but the treatment for the condition at >$500,000/QALY would not be cost-effective even under alternate criteria.

Two studies have assessed the cost-effectiveness of providing ERT, separate from NBS for Pompe disease.^37,38^ In the Netherlands, using a societal perspective, Kanters et al. estimated that alglucosidase alfa treatment of Pompe disease compared to no treatment was greater than €1 million/QALY.^37^ A study from a health sector perspective, estimated a cost of £234,308/QALY in England but did not include outpatient medical costs.^38^ Neither study assessed the cost-effectiveness of NBS for Pompe disease. Our results for the cost-effectiveness of treatment with ERT for clinically-identified infantile-onset Pompe disease appear broadly consistent with these studies.

Our results were not sensitive to the costs of screening. Determining the costs of NBS for a single condition can be difficult if screened for as part of a panel of lysosomal storage disorders. If costs of screening for Pompe disease as part of a panel were just $1, the ICER would still be $333,000/QALY, which is substantially greater than conventional cost-effectiveness thresholds. In the future, better information on costs of screening from NBS programs could improve the accuracy of cost-effectiveness analyses, although that may matter more for conditions with relatively low treatment costs.

In contrast, the results were sensitive to the cost of treatment. The high cost of treatments for rare conditions has been the subject of increasing debate.^39^ Reducing the costs of ERT would result in more favorable cost-effectiveness ratios for NBS. If the cost of alglucosidase alfa were reduced by 80%, the cost-effectiveness ratio for NBS for infantile-onset Pompe disease would fall below $150,000/QALY.

Inclusion of changes in quality of life for family members of a child with infantile-onset Pompe (family spillover effects) did not substantially affect the cost-effectiveness ratios. While the earlier initiation of ERT can improve the quality of life for children with Pompe disease, it does not eliminate spillover; and as lifespan increases, the duration of spillover also increases.

The use of a societal perspective instead of a narrower healthcare sector perspective had a modest effect on the ICER remaining within the same order of magnitude as the ICER using a healthcare sector perspective. The additional costs included in the societal analysis were informal caregiving and patient time costs, which, while substantial, were modest relative to the cost of ERT (Supplemental Table S11, Impact Inventory). Since Pompe disease is not associated with cognitive impairment, special education costs would not be applicable. Although productivity costs are substantial, they are offset by reduced consumption costs; guidance for the inclusion of both categories for pediatric populations is not currently available.

The evidence base for Pompe disease and other rare conditions is scarce. Lack of long-term outcomes data for treated patients required us to rely on expert judgment to estimate health trajectories for treated patients. Evidence on costs of living with Pompe disease was also sparse. Despite the uncertainty in these parameters, our results were robust except to changes in the costs of care for infantile-onset Pompe disease. Due to the complexity of the disease model and the size of the modeled cohort, it was not feasible to conduct probabilistic sensitivity analysis in the study due to computational limitations.

The use of Medicare fee schedules is a commonly-used approach by cost-effective analysts for costing health service utilization.^10^ In recent years, the Centers for Medicare and Medicaid Services has not increased physician fees for many services and this could result in a potential limitation for this study. If costs of medical services are underestimated, this would underestimate the benefits of screening and earlier treatment, yielding less favorable ratios than if costs of medical care were higher. However, since ERT represents the majority of the health care costs for treated individuals, the impact of this limitation would be limited.

Newborn screening for infantile-onset Pompe disease also has implications for identification and treatment of late-onset Pompe disease, but these were not captured in this study to be consistent with the ACHDNC process to evaluate the benefits and harms of conditions that would be addressed during the newborn and childhood ages (typically the first year of life).^3^ The evidence review conducted for the ACHDNC focused on infantile-onset Pompe disease, consistent with the policy question faced by ACHDNC.^6^ Our analysis does include the costs of following probable late-onset cases but does not include any benefits or costs associated with treatment with ERT. One study has found ERT to improve the quality of life of those with late-onset Pompe during the first year of treatment, and to stabilize the quality of life during the subsequent years of treatment ^40^, however treatment protocols for the use of ERT in late-onset patients are still being established. Newer treatment guidelines suggest that early initiation of ERT is beneficial for some patients with late-onset Pompe disease, particularly those with less-advanced disease.^41^ Genetic testing following newborn screening can help identify individuals who are most likely to experience late-onset Pompe and eventually need ERT. Some pathogenic *GAA* variants are strongly correlated with late-onset and not infantile-onset Pompe disease.^42^ Since newborn screening is likely to result in regular clinical follow-up for individuals identified through screening but not diagnosed with infantile-onset Pompe disease, this will likely lead to earlier initiation of ERT for late-onset Pompe disease as a result of newborn screening. As more data become available, future studies should evaluate the cost-effectiveness of early diagnosis and treatment for late-onset Pompe disease. However, because of the cost of monitoring individuals with late-onset Pompe disease over many years and because the benefit of diagnosis may not occur for decades, including late-onset Pompe disease would lead to a less favorable cost-effectiveness assessment. Unfortunately, there are not sufficient data to model the inclusion of late-onset Pompe disease in this microsimulation study.

Newborns with pseudodeficiency may also experience additional costs following identification during NBS even though pseudodeficiency is not associated with any adverse health outcomes. Studies in Canada found modestly higher healthcare utilization by children who screened positive for certain conditions but did not receive a diagnosis.^43,44^ Future studies evaluating long-term outcomes from NBS for Pompe disease could consider collecting data on this group of infants as well.

## Conclusions

Newborn screening for infantile-onset Pompe disease yields considerable health gains and may be cost-effective in the context of rare diseases. As other rare heritable disorders are considered for NBS, this study provides insights into the relative costs and health benefits of screening. First, NBS may be considered cost-effective relative to no screening even when the underlying treatment is not considered cost-effective. At the same time, high-priced treatments may preclude

NBS from being viewed as cost-effective despite substantial health gains. Strategies to reduce the costs of treatment could impact the value of NBS.

## Data Availability

The manuscript and supplementary materials are designed to provide sufficient information for an interested reader to replicate the analysis. The statistical code is available by request from Dr. Prosser.

## Acknowledgements

The authors would like to acknowledge the expert panelists for reviewing assumptions and parameter inputs for the simulation model: Priya Kishnani, MBBS; Deborah Marsden, MD, MBBS; C. Ronald Scott, MD; Olaf Bodamer, MD, PhD; Barry Byrne, MD, PhD; Sharon Kardia, PhD. We would also like to acknowledge the following individuals for providing information on Pompe disease newborn screening: Janice Bach, (Michigan), Patrick Hopkins (Missouri), Tess Rhodes (Illinois), Sarah Bradley (New York), Suzanne Karabin (New Jersey), and Deeksha Bali (Duke University). We would also like to acknowledge Brittany D’Cruz and Nicole Emblad for their research assistance and Rachel Fisher, MS, for her expertise. We are grateful to two anonymous reviewers whose careful review and thoughtful comments have greatly improved this paper.

Financial support for this study was provided by a grant from the Agency for Health Care Research and Quality, R01 HS020644. The funding agreement ensured the authors’ independence in designing the study, interpreting the data, writing, and publishing the report.

## Abbreviations

ACHDNC-: Advisory Committee on Heritable Disorders in Newborns and Children;
ERT-: enzyme replacement therapy;
ICER-: incremental cost-effectiveness ratio;
NBS-: newborn screening;
NICE-: National Institute for Health and Clinical Excellence;
QALYquality-: adjusted life year;
RUSP-: Recommended Uniform Screening Panel

## Disclaimers

The findings and conclusions in this report are those of the authors and do not necessarily represent the official position of the Centers for Disease Control and Prevention. The views expressed herein are solely those of the authors and do not necessarily reflect the views of the Secretary of the United States Department of Health and Human Services or of the individual members of the Secretary’s Advisory Committee on Heritable Disorders in Newborns and Children.

---

**Figure S1.**
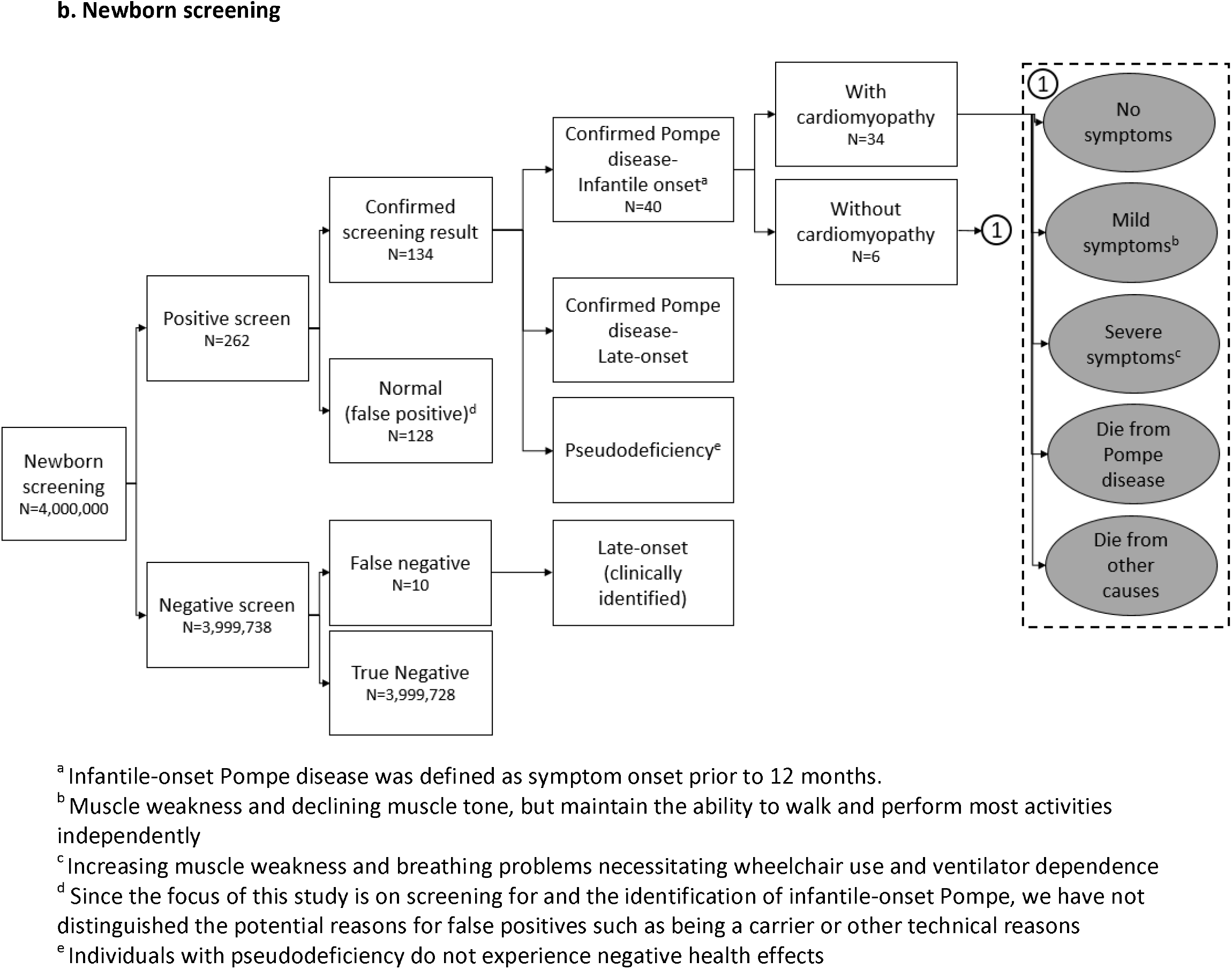
Incremental cost-effectiveness ratio for newborn screening for Pompe disease by cost of alglucosidase alfa (ERT), $/QALY. Abbreviations: ERT, enzyme replacement therapy; QALY, quality adjusted life year; AWP, average wholesale price.

**Figure S2.**
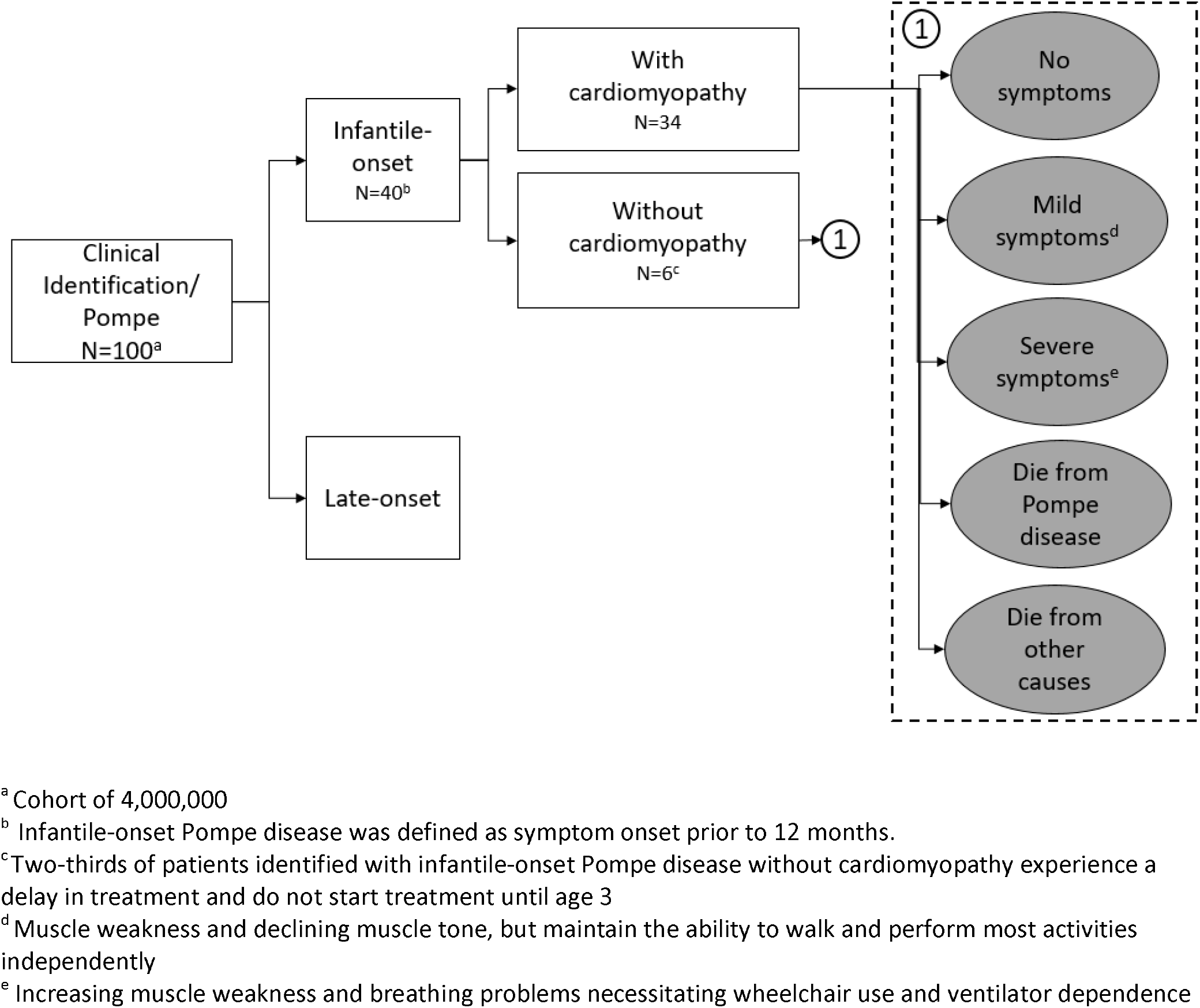
Health state probabilities^a^. a. **Infantile-onset Pompe disease with cardiomyopathy, clinically identified and treated** b. **Infantile-onset Pompe disease without cardiomyopathy, clinically identified and treated** c. **Infantile-onset Pompe disease with cardiomyopathy, newborn screened and treated** d. **Infantile-onset Pompe disease without cardiomyopathy, newborn screened and treated** ^a^Trajectories determined using the transition probabilities outlined in Table S1.

